# Cortical connectivity predicts cognition across time in Parkinson’s disease

**DOI:** 10.1101/2025.05.13.25327462

**Authors:** Hunter P Twedt, Brooke E Yeager, Jacob E Simmering, Jordan L Schultz, Nandakumar S Narayanan

## Abstract

Cognitive symptoms are common in Parkinson’s disease (PD), yet the underlying brain mechanisms remain poorly understood. To address this gap, we investigated the relationship between functional connectivity and cognition at multiple time points using longitudinal functional MRI (fMRI) and cognitive assessments from the Parkinson’s Progression Marker Initiative (PPMI). We calculated resting-state functional connectivity within and between three key cortical brain networks that have been linked with cognitive function in PD: the frontoparietal network (FPN); the salience network (SAL); and the default mode network (DMN). Cognitive function was assessed with the Montreal Cognitive Assessment (MoCA). Linear mixed-effects modeling revealed that decreased FPN-DMN functional connectivity is associated with lower MoCA scores over time. This finding suggests that cortical connectivity is associated with and may contribute to the progression of cognitive symptoms in PD. Our findings advance knowledge about cognitive changes in PD and emphasize the importance of functional brain network architecture.

**Highlights:**

- We studied cortical functional connectivity and cognition in PD over time.
- FPN-DMN functional connectivity robustly predicted cognition at multiple time points.
- These data provide insight into brain networks underlying cognitive progression in PD.

## Introduction

Cognitive impairment in Parkinson’s disease (PD) is an important clinical problem affecting approximately 30% of newly diagnosed patients (Cavanagh et al., 2022; Meireles & Massano, 2012; Narayanan & Albin, 2022). This prevalence increases dramatically with time, with up to 80% of patients experiencing cognitive decline within 20 years of disease onset (Foltynie et al., 2004; Hely et al., 2008). Cognitive symptoms in PD increase the risk of progressing to mild cognitive impairment (PD-MCI) or Parkinson’s disease dementia (PDD) (Lawson et al., 2014; Macleod et al., 2014). Furthermore, cognitive impairment diminishes quality of life and predicts increased mortality (Lawson et al., 2014; Macleod et al., 2014). Despite the immense impact of cognitive symptoms in PD, therapeutic options remain limited due to an incomplete understanding of the underlying brain mechanisms.

Although PD affects several cognitive domains, executive dysfunction involving working memory, attention, and timing is particularly pronounced (Cavanagh et al., 2022; Diamond, 2013; Gilbert & Burgess, 2008; Kudlicka et al., 2011; Parker et al., 2013; Zgaljardic et al., 2003). These functions rely on canonical cortical brain networks, such as the frontoparietal network (FPN), the salience network (SAL), and the default mode network (DMN) (Bressler & Menon, 2010; M. W. Cole et al., 2014; Seeley, 2019; Sridharan et al., 2008). Resting-state functional magnetic resonance imaging (rs-fMRI) work by our group and others has implicated these networks in cognitive dysfunction in PD. Specifically, decreased inter-network functional connectivity between the FPN-DMN and the SAL-DMN was associated with impaired cognition (Aracil-Bolaños et al., 2019; Galvez et al., 2024; Putcha et al., 2015, 2016; Yeager et al., 2024). A limitation of this past work is that relationships between network architecture and cognition were evaluated at a single time point; however, changes in the functional connectivity of these networks over time may be associated with changes in cognitive function in PD.

We investigated this question using longitudinal data from the Parkinson’s Progression Marker Initiative (PPMI) database (Marek et al., 2011). We investigated the relationship between network functional connectivity (within and between the FPN, SAL, and DMN; based on rs-fMRI scans) and cognition (assessed using the Montreal Cognitive Assessment (MoCA)), both of which were measured three times over an average of 3 years. In our longitudinal analysis we observed that decreased FPN-DMN functional connectivity is associated with impaired cognitive function in patients with PD. These findings provide insight into how the progressive nature of PD affects brain networks.

## Methods

### 2.1. Study dataset and participants

To investigate the longitudinal functional network correlates associated with cognitive impairment in PD, we analyzed rs-fMRI data from people with PD included in the publicly available Parkinson’s Progression Markers Initiative (PPMI) database (https://www.ppmi-info.org/access-data-specimens/download-data; Marek et al., 2011). The PPMI provides an open-access, de-identified dataset comprising clinical and imaging data from over 850 patients with PD across 12 countries. From this dataset, we identified 27 patients with PD who had at least three available structural and functional MRI scans and MoCA scores collected at least 10 months apart. For each patient, neuroimaging, neuropsychological, and clinical data have been collected for over 3 years on average, facilitating comprehensive and longitudinal studies. Consistent with the PPMI protocol, identifying details, such as scanning site information, are de-identified. Demographic characteristics of the sample are summarized in Table 1. All participants in our PPMI sample were Caucasian/White. Ethical approval for the study was granted by the Institutional Review Board (IRB) of all contributing sites, and all participants provided written informed consent prior to enrollment in the PPMI study. The University of Iowa IRB determined that secondary analysis of public datasets like PPMI are IRB exempt.

**Table 1.**
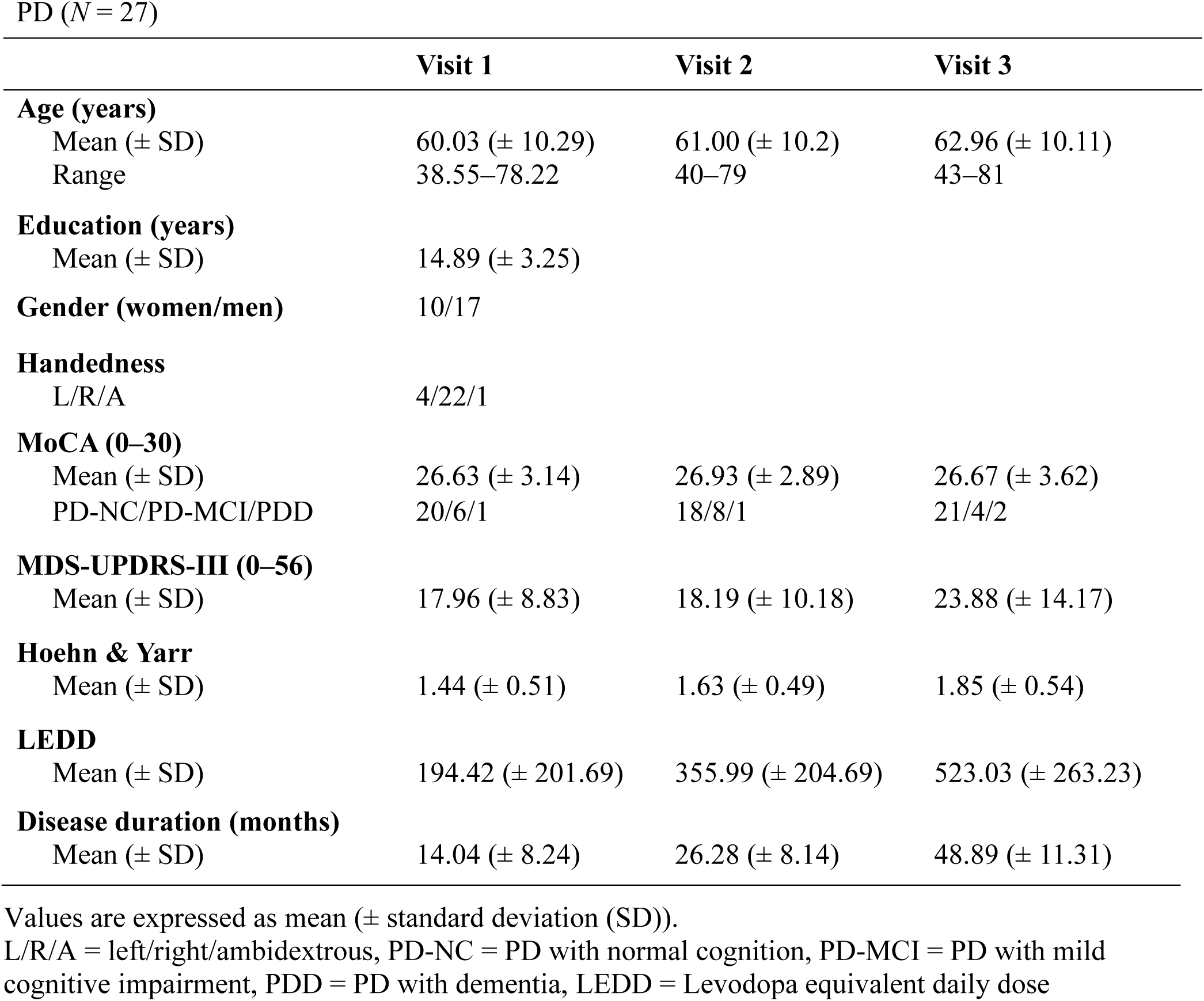
Demographic and clinical data of study population.

### 2.2. Clinical assessments

We identified relevant clinical assessments of motor and non-motor symptoms from the PPMI dataset to use in our analyses. Each clinical assessment described here was collected on the same visit dates as each of the participants three MRI scans. Results from the Montreal Cognitive Assessment (MoCA) were employed as a continuous measure of global cognitive function within our PPMI sample. The MoCA is a widely recognized and validated tool for assessing cognitive abilities in the context of normal aging (Bernier et al., 2023) and in individuals with neurological disorders (Dalrymple-Alford et al., 2010; Freitas et al., 2013; Gill et al., 2008; Nasreddine et al., 2005). Although it was designed as a screening instrument and is not as detailed as in-depth neuropsychological testing, it is widely used in clinical practice (Litvan et al., 2012), and it accurately measures cognitive impairment in PD with very high sensitivity (90%) and specificity (75%) (R. C. Cole et al., 2023; Dalrymple-Alford et al., 2010; Hoops et al., 2009; Singh et al., 2018, 2021). In our PPMI sample, participants with a MoCA score of 26–30 were considered cognitively normal (PD-NC), those with a MoCA score of 22–25 were considered to have mild cognitive impairment (PD-MCI), and those with a MoCA score of <22 were considered to have dementia (PDD). Motor symptom severity was assessed using scores from the Movement Disorder Society-Unified Parkinson’s Disease Rating Scale-Part III (MDS-UPDRS-III), and functional disability was assessed using Hoehn and Yahr staging. For each participant, disease duration was calculated in months from the date of diagnosis to the participant’s fMRI session dates. The sum of the doses of each patient’s parkinsonian medications at each PPMI visit was calculated and is reported as the Levodopa equivalent daily dose (LEDD) (Julien et al., 2021).

### 2.3. MRI acquisition and preprocessing

All participants underwent standardized MRI scanning using a 3T Siemens Trio Tim scanner at one of nine institutions in the United States or Europe. Detailed scanning protocols are described in the PPMI MRI operations manual available at https://www.ppmi-info.org/. T1-weighted structural images were acquired with the following parameters: repetition time (TR) = 2300 ms; echo time (TE) = 2.98 ms; flip angle (FA) = 9°; and voxel size = 1 mm^3^. rs-fMRI scans included 210 volumes with parameters of TR = 2400 ms, TE = 25 ms, FA = 80°, and voxel size = 3.3 mm^3^. The rs-fMRI sequence had a total acquisition time of 8 minutes and 24 seconds, according to the PPMI MRI operations manual. Participants were instructed to rest quietly with their eyes open, to clear their mind, and to not fall asleep; they were also reminded to minimize all movements during the scans. Both T1-weighted structural and rs-fMRI datasets were downloaded and processed using the CONN toolbox (version 21a), an open-source MATLAB Statistical Parametric Mapping (SPM)-based software package (Whitfield-Gabrieli & Nieto-Castanon, 2012). The default preprocessing pipeline for volume-based analyses in CONN was applied, followed by its standard denoising procedures. The preprocessing steps included motion correction (realignment and unwarp); slice-timing correction; and outlier detection, using artifact detection tools (ART) to flag frames with framewise displacement exceeding 1 mm. Data were normalized to MNI space and smoothed using a 6-mm full-width-at-half-maximum (FWHM) Gaussian kernel. Additionally, T1-weighted images were segmented into gray matter, white matter, and cerebrospinal fluid (CSF). We included a whole-brain mask region of interest (ROI) as an additional confound to account for undesired global noise. The denoising pipeline was then implemented to further reduce confounding influences through linear regression, followed by the application of a temporal band-pass filter (0.01–0.08 Hz; He et al., 2008; Schölvinck et al., 2010). Confounding regressors included three translational and three rotational parameters, as well as five temporal derivatives from the white matter and CSF. The first derivative represented the mean blood-oxygen-level-dependent (BOLD) signal, while the remaining four derivatives were derived from a principal component analysis of the covariance within the orthogonal subspace to the mean signal and other nuisance effects (Chai et al., 2012; Nieto-Castanon, 2020).

### 2.4. Functional connectivity analyses

We performed seed-based, resting-state functional connectivity analysis using the default weighted general linear model implemented in the CONN toolbox, as in our past work (Yeager et al., 2024). Both seed-to-voxel and ROI-to-ROI connectivity analyses were utilized to assess functional connectivity patterns. For each participant, seed-to-voxel and ROI-to-ROI Pearson’s correlation connectivity maps were generated that represent the temporal correlations of BOLD activity between ROIs. These first-level correlations were Fisher r-to-z transformed to enhance normality assumptions and exported as subject-level z-maps. Second-level analyses were performed using MATLAB (version R2022b) and R (version 4.3.1) to evaluate group differences. To quantify intra-network connectivity, Fisher-transformed z-values of all ROI-to-ROI pairs within each network were averaged. For inter-network connectivity, mean z-values were calculated for all ROI-to-ROI connections between two networks. Network ROIs provided by the CONN toolbox were informed by independent component analysis of the Human Connectome Project dataset (*N* = 497) (Whitfield-Gabrieli & Nieto-Castanon, 2012).

We focused on three canonical cortical networks that are frequently implicated in the literature on rs-fMRI functional connectivity analyses: the frontoparietal network (FPN); the salience network (SAL); and the default mode network (DMN). These networks were selected based on prior hypotheses suggesting that their functional connectivity is associated with cognitive impairment in PD (Aracil-Bolaños et al., 2019; Baggio et al., 2014, 2015; B. Chen et al., 2015; L. Chen et al., 2022; Galvez et al., 2024; Goulden et al., 2014; Lewis et al., 2012; Lucas-Jiménez et al., 2016; Putcha et al., 2015, 2016). A detailed description of each network’s composition with x, y, z coordinates in MNI space is presented in Table 2.

**Table 2.** CONN toolbox network region of interest (ROI) definitions.

| Network name | Seed | *x* | *y* | *z* |
| --- | --- | --- | --- | --- |
| Frontoparietal network (FPN) | Lateral prefrontal cortex (L) | -43 | 33 | 28 |
|  | Lateral prefrontal cortex (R) | 41 | 38 | 30 |
|  | Posterior parietal cortex (L) | -46 | -58 | 49 |
|  | Posterior parietal cortex (R) | 52 | -52 | 45 |
| Salience network (SAL) | Mid-cingulate cortex | 0 | 22 | 35 |
|  | Anterior insula (L) | -44 | 13 | 1 |
|  | Anterior insula (R) | 47 | 14 | 0 |
|  | Rostral prefrontal cortex (L) | -35 | 45 | 27 |
|  | Rostral prefrontal cortex (R) | 32 | 46 | 27 |
|  | Supramarginal gyrus (L) | -60 | -39 | 31 |
|  | Supramarginal gyrus (R) | 62 | -35 | 32 |
| Default mode network (DMN) | Medial prefrontal cortex | 1 | 55 | -3 |
|  | Lateral parietal cortex (R) | 47 | -67 | 29 |
|  | Posterior cingulate cortex | 1 | -61 | 38 |
|  | Lateral parietal cortex (L) | -39 | -77 | 33 |

### 2.5. Statistical analyses

We harnessed linear mixed-effects modeling to evaluate the relationship between functional connectivity and cognition at three time points over a period of ∼3 years on average. As in our prior work, functional connectivity was the predictor (independent variable) and MoCA scores were considered the outcome (dependent variable). To identify potential covariates, we estimated associations between baseline clinical assessments and demographic variables with MoCA scores using Spearman’s correlation. Factors with p values ≤0.05 were determined to be significantly associated with MoCA scores and were thus included in our linear mixed-effect models. Given that longitudinal data include repeated measures for each participant and may cause clustering of observations among patients, we used a mixed-effects linear model. Our primary model regressed MoCA score on functional connectivity measure, time since baseline, age, education, MDS-UDPRS III score, total LEDD, and a random intercept for each study participant, with three observations per patient corresponding to the three independent time points. LEDD was included as a covariate in our model because previous work suggests that parkinsonian medication leads to dopaminergic modulation of functional brain networks (Berman et al., 2016; Day et al., 2019; Kelly et al., 2009; Krajcovicova et al., 2012; Yang et al., 2016), and dopaminergic medication may have variable effects on cognition (Cools, 2006; Ikeda et al., 2017; Roy et al., 2018). We considered *p* values ≤0.05 as statistically significant. All analyses were conducted in R (version 4.4.2), using the *lmer* package for linear mixed-effects modeling (Bates et al., 2015) and the *lmerTest* package for hypothesis testing (Kuznetsova et al., 2017). In these models, Satterthwaite’s method was used to generate the *p* values. Data and analysis code are publicly accessible at narayanan.lab.uiowa.edu.

## Results

### Cohort Characteristics

Demographic, neuropsychological, and clinical characteristics of our sample are summarized in Table 1. The average time between the first and second MRI scanning sessions (Visit 1 and Visit 2) was 11.7 months (SD: 0.82), and the average time between the first and third scanning sessions (Visit 1 and Visit 3) was 34.3 months (SD: 6.71). When we evaluated whether neuropsychological and clinical measures changed over time in our sample, we found that MoCA scores (Figure 1) did not reliably change (*p* = 0.87). There was a trend of increased motor symptom severity, as measured with the MDS-UPDRS-III, although this trend was not statistically significant (*p* = 0.07). However, we found that participants in our PPMI dataset experienced functional decline, with Hoehn and Yahr stages increasing significantly over time (*p* = 0.003), from 1.44 (SD: 0.51) at the first visit to 1.85 (SD: 0.54) at the third visit. Participants also had increased doses of parkinsonian medication over time (LEDD; *p* < 0.001). Additionally, we identified several variables that had significant univariate associations with MoCA scores, including age (*r* = -0.38, *p* = 0.0004), education (*r* = 0.26, *p* = 0.02), and MDS-UPDRS-III (*r* = -0.28, *p* = 0.01; Table 3). These variables were included as covariates in our linear mixed-effects models.

**Figure 1.**
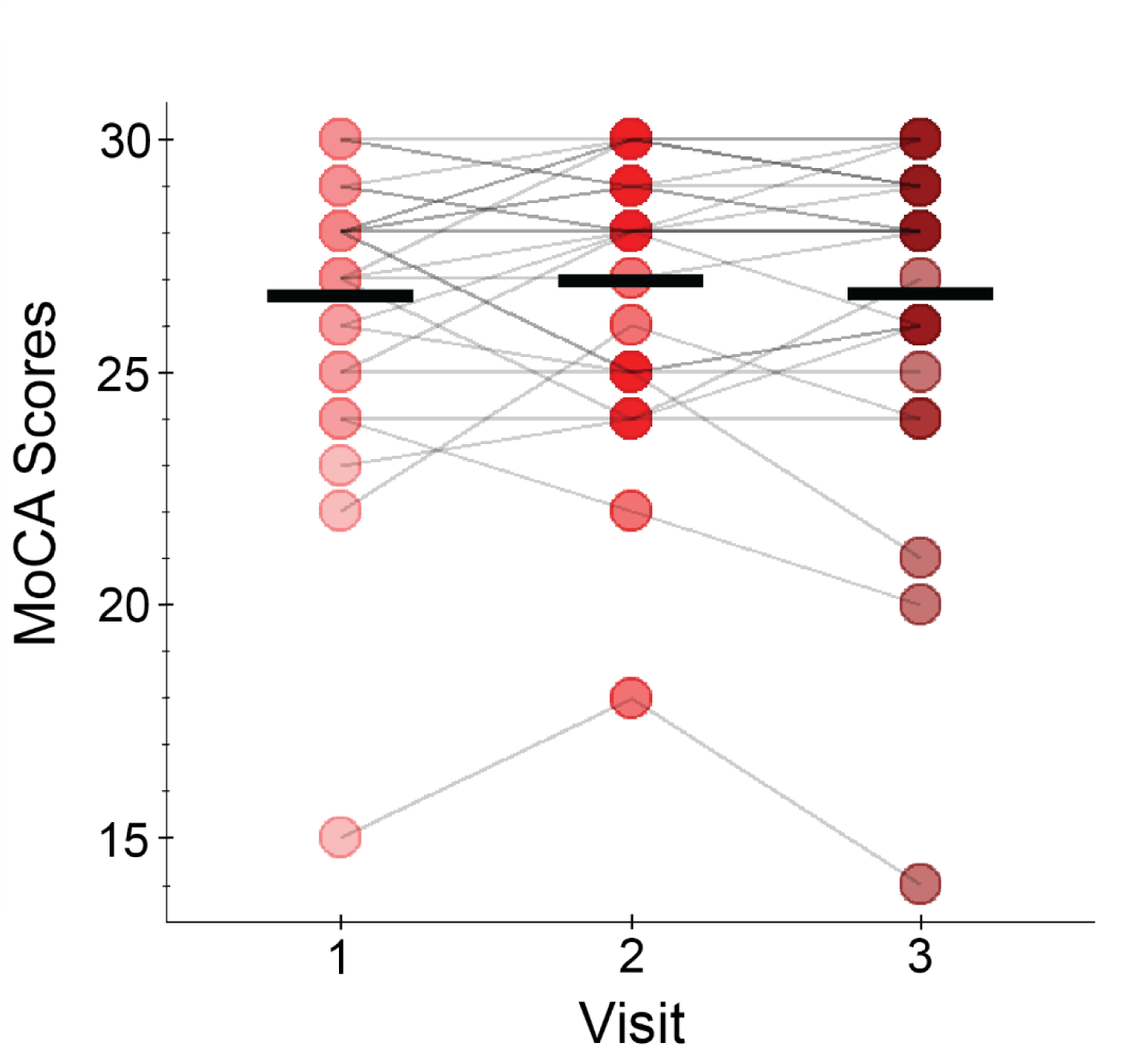
In patients with PD in our PPMI dataset, MoCA scores from Visit 1 to Visit 3 (an average of ∼3 years) did not significantly change. Black bars represent mean MoCA scores. Gray lines connect data from individual participants across visits.

**Table 3.** Correlation matrix of study variables.

|  | **MoCA** | **Age** | **Education** | **MDS-**<br>**UPDRS-III** | **Hoehn &**<br>**Yarr** | **Disease**<br>**duration** |
| --- | --- | --- | --- | --- | --- | --- |
| **MoCA** | - |  |  |  |  |  |
| **Age** | -0.38<br>[-0.51, -0.12] | - |  |  |  |  |
| **Education** | 0.26<br>[-0.17, 0.27] | -0.24<br>[-0.34, 0.10] | - |  |  |  |
| **MDS-**<br>**UPDRS-**<br>**III** | -0.28<br>[-0.58, -0.22] | 0.35<br>[0.19, 0.56] | 0.03<br>[-0.13, 0.31] | - |  |  |
| **Hoehn &**<br>**Yarr** | -0.14<br>[-0.43, -0.01] | 0.12<br>[-0.09, 0.35] | 0.34<br>[0.15, 0.53] | 0.58<br>[0.45, 0.73] | - |  |
| **Disease**<br>**duration** | -0.03<br>[-0.42, 0.001] | 0.31<br>[0.08, 0.48] | 0.05<br>[-0.20, 0.24] | 0.20<br>[0.10, 0.50] | 0.27<br>[0.12, 0.51] | - |
| **LEDD** | -0.14<br>[-0.42, -0.0005] | 0.26<br>[0.03, 0.44] | -0.09<br>[-0.29, 0.14] | 0.01<br>[-0.17, 0.27] | 0.09<br>[-0.12, 0.31] | 0.67<br>[0.54, 0.78] |

Values are given as correlation estimates [95% confidence intervals].

### Functional Connectivity

We found no reliable impact of within-network functional connectivity of the FPN, SAL, and DMN on cognition, as measured by the MoCA (Table 4). Next, we examined the effect of inter-network functional connectivity (FPN-DMN, SAL-DMN, FPN-SAL) on cognition. When controlling for significant univariate predictors of MoCA scores, we found that increased FPN-DMN functional connectivity was significantly associated with increased MoCA scores at each of the three time points (*β* = 3.07, 95% CI [0.45, 5.73], suggesting that lower FPN-DMN functional connectivity predicted cognitive progression over time (Figure 2). We also found a similar but nonsignificant association between SAL-DMN functional connectivity and cognitive progression (*β* = 3.22, 95% CI [-0.08, 6.53]; Figure S1).

**Figure 2.**
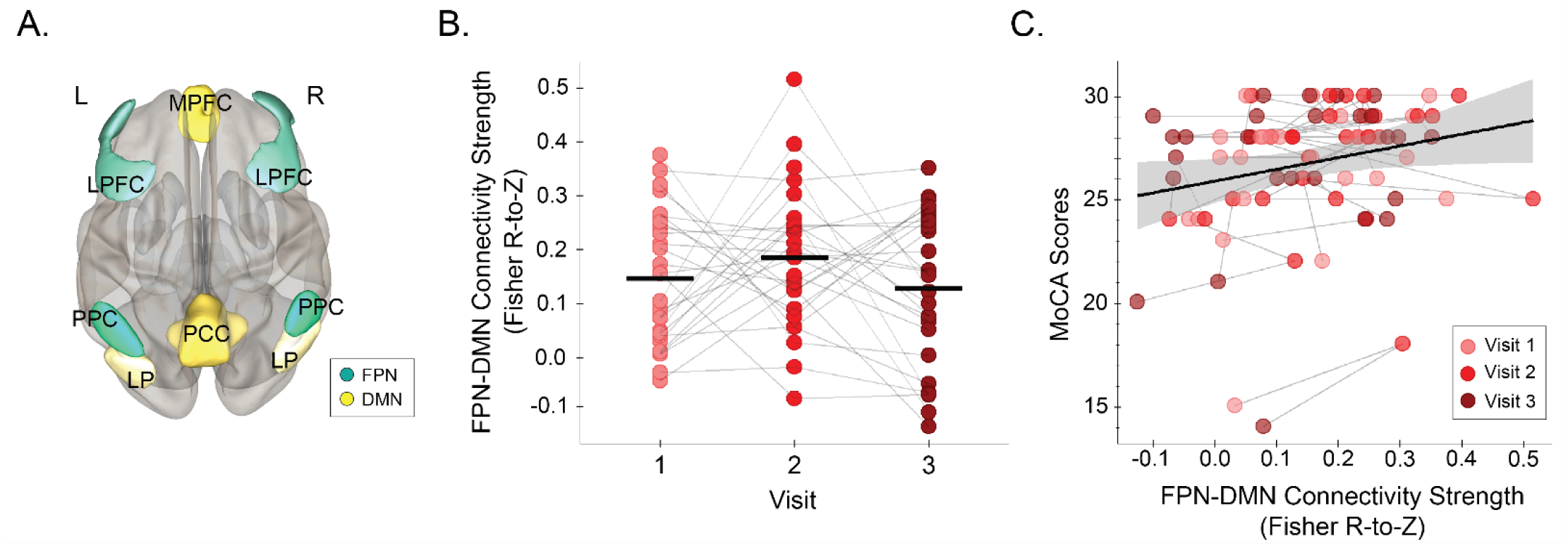
FPN-DMN inter-network functional connectivity (*N* = 27). A) 3D-rendered display of FPN-DMN regions of interest (ROIs) from the superior view of the brain. B) Differences in FPN-DMN inter-network functional connectivity across Visits 1–3 for each participant, over a total of ∼3 years on average. FPN-DMN functional connectivity did not significantly differ across Visits 1–3 (*p* = 0.36). C) Scatterplot showing the relationship between FPN-DMN functional connectivity (Fisher R-to-Z values) and MoCA scores for each participant across Visits 1–3. Data for each participant are plotted individually. The colors of the datapoints (pink, red, and maroon) represent Visits 1, 2, and 3, respectively; a thin gray line connects the datapoints for each participant. The black line represents our overall linear regression model regressing MoCA scores on FPN-DMN functional connectivity, with adjustments for time, age, education, MDS-UPDRS-III, LEDD, and a random intercept by participant. Gray bands denote the 95% confidence interval. *LPFC = lateral prefrontal cortex, PPC = posterior parietal cortex, MPFC = medial prefrontal cortex, PCC = posterior cingulate cortex, LP = lateral parietal cortex*.

**Table 4.** Linear model estimates of functional connectivity relationships with PD symptoms.

|  | MoCA |  |  |  | MDS-UPDRS-III |  |  |  |
| --- | --- | --- | --- | --- | --- | --- | --- | --- |
|  | *Beta* | *t-stat (df)* | *95% CI* | *p* | *Beta* | *t-stat (df)* | *95% CI* | *p* |
| **FPN** | -0.51 | -0.42 (58.26) | [-2.78, 2.02] | 0.67 | 7.65 | 1.40 (66.61) | [-3.98, 18.01] | 0.17 |
| **SAL** | 1.64 | 0.92 (53.01) | [-1.73, 5.26] | 0.36 | 8.53 | 1.00 (61.53) | [-8.27, 24.81] | 0.32 |
| **DMN** | 0.04 | 0.03 (64.71) | [-2.75, 2.86] | 0.98 | -4.71 | -0.72 (73.61) | [-17.70,7.67] | 0.48 |
| ***FPN-DMN\**** | *3.07* | *2.24 (51.23)* | *[0.45, 5.73]* | *0.03* | *-8.33* | *-1.21 (59.47)* | *[-21.62, 4.88]* | *0.23* |
| **FPN-SAL** | -1.49 | -0.74 (55.72) | [-5.27, 2.45] | 0.46 | 3.02 | 0.32 (68.28) | [-15.48, 21.37] | 0.75 |
| **SAL-DMN~** | 3.22 | 1.87 (56.49) | [-0.08, 6.53] | 0.07 | -0.52 | -0.06 (69.46) | [-16.45, 15.96] | 0.95 |

df = degrees of freedom, CI = confidence interval, \* = $p < 0.05$ , ~ = $p > 0.05$ and $<0.10$

Notably, neither FPN-DMN (*β* = -8.33, 95% CI [-21.62, 4.88]) nor SAL-DMN (*β* = -0.52, 95% CI [-16.45, 15.96]) functional connectivity were associated with changes in MDS-UPDRS-III scores over time. The specificity of the relationship between inter-network functional connectivity and cognition but not motor symptoms suggests that an association is not easily explained by greater PD disease burden or motor impairment. We did not find other reliable longitudinal inter-network associations with MoCA scores (Table 4). Taken together, these data provide insight into the functional network alterations that may predict cognitive progression in PD.

## Discussion

We tested the hypothesis that functional connectivity is associated with cognition over time in PD. We focused on cognitive brain networks known to be disrupted in PD (FPN, SAL, DMN and cognitive function as measured by the MoCA, a widely-used measure of cognitive function (Amboni et al., 2015; Aracil-Bolaños et al., 2019; Galvez et al., 2024; Putcha et al., 2015, 2016; Yeager et al., 2024). We found that cognition in PD is associated with inter-network functional connectivity. Specifically, we showed a significant positive relationship between FPN-DMN functional connectivity and MoCA scores, indicating that a reduction in functional connectivity is associated with a lower average cognitive function, after adjustment for confounds and accounting for repeated measures. FPN-DMN functional connectivity had a specific relationship to cognitive function as there was no reliable relationship between inter-network functional connectivity and motor function as measured by the MDS-UPDRS-III. These data provide novel insight about the relationship between functional brain networks and cognition in PD and indicate that brain network changes are associated with cognitive decline in PD.

This relationship between FPN-DMN functional connectivity and cognition aligns with previous studies indicating that the FPN plays a crucial role in executive functions such as inhibition, working memory, cognitive flexibility, and timing, which are often impaired in PD patients (Amboni et al., 2015; Aracil-Bolaños et al., 2019; Putcha et al., 2015, 2016). The DMN has also been previously identified as a major contributor to cognition in PD (Gorges et al., 2015), a finding supported by our current results. Moreover, our prior work found that FPN-DMN connectivity predicted cognitive impairments in PD at a single time point (Yeager et al., 2024). Here, we expand on that finding by demonstrating that longitudinal measures of inter-network functional connectivity are associated with changes in cognition in patients with PD over time. Alterations in the posterior parietal nodes of the FPN and DMN, which are regions vulnerable to metabolic changes in PD, may underlie the cognitive deficits observed in patients (Firbank et al., 2017; Isaias et al., 2020).

Our work has several limitations. First, in this population changes in MoCA scores and changes in FPN-DMN or SAL-DMN functional connectivity were relatively slow; we did not observe significant group trends in MoCA scores or in FPN-DMN or SAL-DMN functional connectivity over the 3 years of data available. However, our analysis does find that the variability between these measures over time is correlated with declines in MoCA being associated with changes in functional connectivity. An analysis of data with a longer duration of follow-up—and more within-person variability in cognitive and functional connectivity measures—would provide greater power to estimate these associations. Second, our sample included only a small number of participants with MoCA scores that were low enough to be classified as having PD-MCI (defined as a score of ≥22 and ≤25) or PDD, defined as a score of <22 (Table 1), limiting our ability to investigate the neural circuitry associated with more severe cognitive decline. Third, we selected the MoCA because it is a widely used and validated global measure of cognitive function, but changes in functional connectivity may impact specific cognitive domains differently. Since PD has multiple phenotypes of cognitive impairment (e.g., frontostriatal, posterior cortical, and mixed subtypes (Kehagia et al., 2012; Summers et al., 2024)), future studies should administer more intensive cognitive testing that provides cognitive-domain-specific measures (Dalrymple-Alford et al., 2010). In addition, these studies should focus on a richer classification of the network substrates associated with each cognitive subtype (Chung et al., 2019; Devignes et al., 2022; Lang et al., 2019). Fourth, our analyses of network connectivity relied on ROIs derived from a normative sample within the Human Connectome Project (Nieto-Castañón & Fedorenko, 2012). We assume that the network ROIs generalize to patients with PD; however, this has not yet been validated. Future work should also aim to develop a diverse PD-specific network parcellation to refine the Human Connectome Project ROIs and validate ROIs in a PD population. Fifth, the PPMI database does not have longitudinal fMRI data for their sample of healthy controls. Thus, we were unable to compare our assessment of functional connectivity relationships on cognition in PD samples over time with healthy adults, making us unable to infer whether our results are PD-specific or reflect aging more generally. Indeed, in elderly individuals without neurological disease, decreased SAL connectivity is related to cognitive decline (Onoda et al., 2012). However, whether SAL dynamics are disrupted to a greater extent in patients with PD compared with healthy elderly adults is not known. Therefore, future work is needed to compare longitudinal network data between patients with PD and healthy elderly adults to accurately define such relationships.

In summary, our current study provides novel insights into the functional neural correlates of cognitive decline in PD and emphasizes the importance of inter-network architecture in supporting cognition. We find that cortical network connectivity between the FPN-DMN was reliably related to cognition in patients with PD over time. These results highlight the importance of large-scale network dysfunction in PD-related cognitive decline and provide a foundation for future research aimed at identifying biomarkers for cognitive dysfunction in PD and other neurodegenerative diseases.

## Data Availability

All data are available online at Open Neuro (https://openneuro.org/datasets/ds006224).

https://www.ppmi-info.org/access-data-specimens/download-data

## Contributors

HPT and BEY conceptualized and designed the study. HPT and BEY conducted the analyses. HPT, BEY, JES, JLS, and NSN interpreted the data. HPT wrote the original draft of the manuscript. HPT, BEY, JES, JLS, and NSN edited and revised the manuscript.

## Ethics approval

The PPMI study was approved by the local Institutional Review Board of each participating institution (a full list is available at https://www.ppmi-info.org/about-ppmi/ppmi-clinical-sites). Written informed consent was obtained from each participant at enrollment, in accordance with the Declaration of Helsinki. All methods were performed in accordance with relevant guidelines and regulations.

## CRediT authorship contribution statement

Hunter P. Twedt: Writing—Original draft, Writing— Review & editing, Visualization, Methodology, Investigation, Formal analysis, Conceptualization. Brooke E. Yeager: Writing—Review & editing, Visualization, Methodology, Investigation, Formal analysis, Conceptualization. Jacob Simmering: Writing—Review & editing, Formal analysis. Jordan L Schultz: Writing—Review & editing. Nandakumar S. Narayanan: Writing—Review & editing, Supervision, Funding acquisition, Formal analysis.

## Data availability

Data are available upon reasonable request. All data and code are available at narayanan.lab.uiowa.edu.

## Acknowledgments

We want to thank Heather Widmayer with the Scientific Editing and Research Communication Core at the University of Iowa Carver College of Medicine for thoroughly editing our manuscript.

Data used in the preparation of this article were obtained June 25, 2021, from the Parkinson’s Progression Markers Initiative (PPMI) database (www.ppmi-info.org/access-data-specimens/download-data). For up-to-date information on the study, visit www.ppmi-info.org. **Funding**: PPMI—a public– private partnership—is funded by the Michael J. Fox Foundation for Parkinson’s Research funding partners, including 4D Pharma, Abbvie, AcureX, Allergan, Amathus Therapeutics, Aligning Science Across Parkinson’s, AskBio, Avid Radiopharmaceuticals, BIAL, Biogen, Biohaven, BioLegend, BlueRock Therapeutics, Bristol-Myers Squibb, Calico Labs, Celgene, Cerevel Therapeutics, Coave Therapeutics, DaCapo Brainscience, Denali, Edmond J. Safra Foundation, Eli Lilly, Gain Therapeutics, GE HealthCare, Genentech, GSK, Golub Capital, Handl Therapeutics, Insitro, Janssen Neuroscience, Lundbeck, Merck, Meso Scale Discovery, Mission Therapeutics, Neurocrine Biosciences, Pfizer, Piramal, Prevail Therapeutics, Roche, Sanofi, Servier, Sun Pharma Advanced Research Company, Takeda, Teva, UCB, Vanqua Bio, Verily, Voyager Therapeutics, the Weston Family Foundation, and Yumanity Therapeutics.

