## Supplemental Figure 1 for "Cortical connectivity predicts cognition across time in Parkinson’s disease"

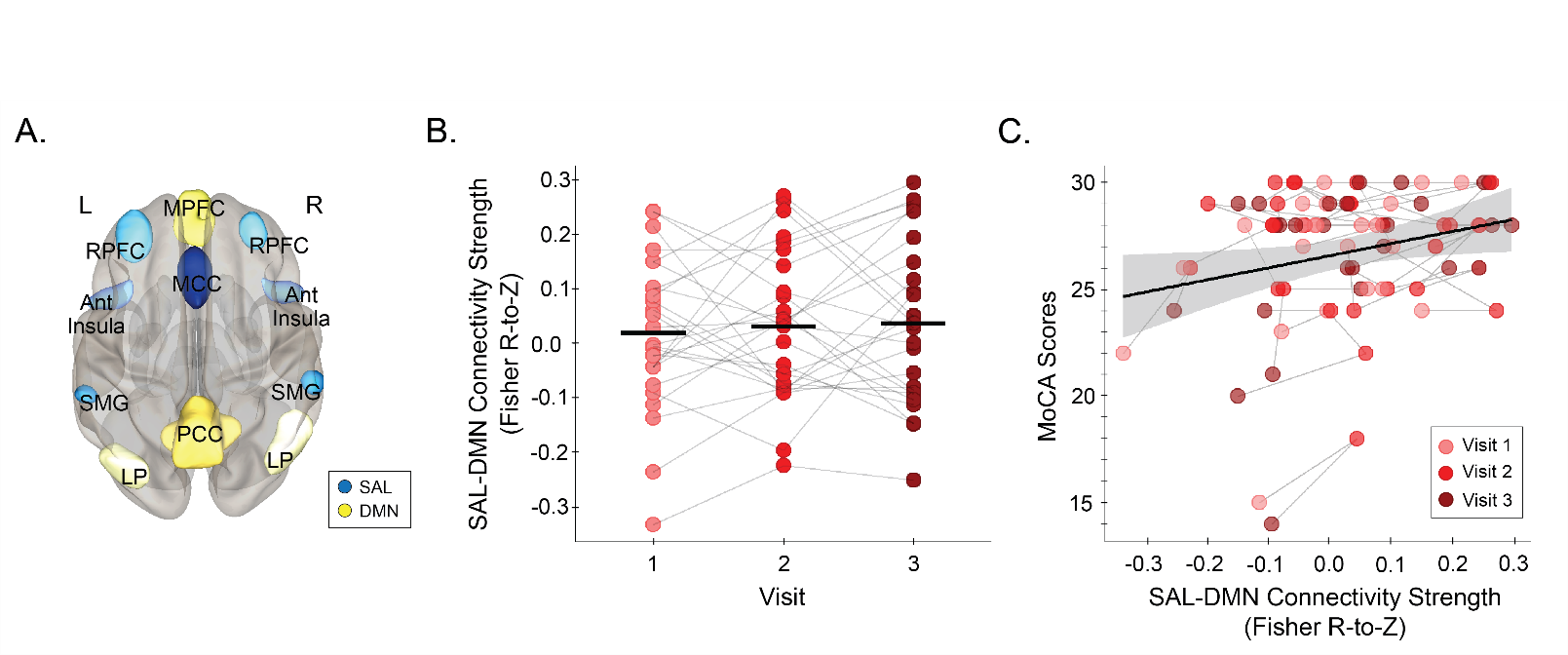
**Figure S1.** SAL-DMN inter-network functional connectivity (*N* = 27). A) 3D-rendered display of SAL-DMN regions of interest (ROIs) from the superior view of the brain. B) Differences in SAL-DMN inter-network functional connectivity across Visits 1–3 for each participant, over a total of ~3 years on average. SAL-DMN functional connectivity did not significantly differ across Visits 1–3 (*p* = 0.57). C) Scatterplot showing the relationship between SAL-DMN functional connectivity (Fisher R-to-Z values) and MoCA scores for each participant across Visits 1–3. Data for each participant are plotted individually. The colors of the datapoints (pink, red, and maroon) represent Visits 1, 2, and 3; a thin gray line connects the datapoints for each participant. The black line represents our overall linear regression model regressing MoCA score on SAL-DMN functional connectivity, with adjustments for time, age, education, MDS-UPDRS-III, LEDD, and a random intercept by participant. Gray bands denote the 95% confidence interval. *MPFC = medial prefrontal cortex, PCC = posterior cingulate cortex, LP = lateral parietal cortex, RPFC = rostral prefrontal cortex, MCC = mid-cingulate cortex, Ant Insula = Anterior Insula, SMG = superior marginal gyrus.*
